# Machine learning models for predicting severe COVID-19 outcomes in hospitals

**DOI:** 10.1101/2022.10.28.22281646

**Authors:** Philipp Wendland, Vanessa Schmitt, Jörg Zimmermann, Lukas Häger, Siri Göpel, Christof Schenkel-Häger, Maik Kschischo

**Affiliations:** University of Applied Sciences Koblenz, Department of Mathematics and Technology, Remagen, DE; University Clinic Tübingen, Department of Internal Medicine 1, Tübingen, DE; University of Applied Sciences Koblenz, Department of Economics and Social Care, Remagen, DE

**Keywords:** Covid-19, Machine Learning, Prediction, Disease Progression, Laboratory values, Precision medicine

## Abstract

The aim of this observational retrospective study is to improve early risk stratification of hospitalized Covid-19 patients by predicting in-hospital mortality, transfer to intensive care unit (ICU) and mechanical ventilation from electronic health record data of the first 24 hours after admission. Our machine learning model predicts in-hospital mortality (AUC=0.918), transfer to ICU (AUC=0.821) and the need for mechanical ventilation (AUC=0.654) from a few laboratory data of the first 24 hours after admission. Models based on dichotomous features indicating whether a laboratory value exceeds or falls below a threshold perform nearly as good as models based on numerical features. We devise completely data-driven and interpretable machine-learning models for the prediction of in-hospital mortality, transfer to ICU and mechanical ventilation for hospitalized Covid-19 patients within 24 hours after admission. Numerical values of CRP and blood sugar and dichotomous indicators for increased partial thromboplastin time (PTT) and glutamic oxaloacetic transaminase (GOT) are amongst the best predictors.

## 1. Introduction

Beginning in late 2019 and lasting until now SARS-CoV-2 manifested as Covid-19 spread all over the world and caused a worldwide pandemic. Infected patients develop a variety of disease symptoms and differences in the hemogram resulting in a wide range of disease severity from mild symptoms not requiring any medical intervention to mechanical ventilation or a transfer to intensive care unit (ICU) or even death [1–3]]. Several drugs for Covid-19 treatment have been developed since the beginning of the pandemic and most of them are linked to different disease stages. For example, hospitalized patients with severe symptoms can be treated with Remdesivir and Dexamethason, wheareas antibody-based therapy had to be administered at an early disease stage before a patient has developed severe symptoms [4,5].

For optimal patient care and treatment in hospitals it is very important to detect patients with bad prospective disease progression early and to devise reliable prediction models which can easily be applied in daily clinical practice. Such an early stratification of patients can already be used for a decision whether ambulant treatment is sufficient or whether hospital admittance is advisable. More elaborate and costly diagnosis can be targeted at high risk patients, e.g. a computertomography of the thorax instead of conventional X-ray or a more comprehensive blood count. High risk patients can be more intensively monitored, e.g. by more frequent laboratory tests, oxygen saturation measurements and blood gas analysis leading to an earlier detection of a worsening of the disease state, so that a medical intervention can be initiated by physicians. In summary, the early identification of high risk patients can both improve patient outcome and alleviate the overwhelming pressure on hospitals experienced during the last and possible future pandemics.

Many existing predictive models of severe Covid-19 disease progression are based on data from tertiary care hospitals like university hospitals or from clinical study data repositories. Many scoring models incorporate non-standard laboratory values or are only based on diagnoses, which renders their widespread application in daily clinical practice difficult [6–8]. Here we present personalized and completely data-driven machine-learning models for the prediction of (i) in-hospital mortality, (ii) transfer to ICU and (iii) mechanical ventilation of hospitalized Covid-19 patients. Our models use standard clinical laboratory data from hospitals of medium level of care measured during clinical routine in combination with biological sex and age as covariates. Our purely data-driven approach avoids potential bias or the pure reproduction of well-known results [9] and is an important addition to the landscape of expert knowledge-based Covid-19 risk scores [10–13]. In contrast to previous approaches [14], we explored a large space of potential predictors.

We also present simplified models using only dichotomous predictors indicating whether a laboratory value is below or above reference threshold. These might better reflect the daily clinical practice than a complex combination of numerical features. In addition, we report a comprehensive analysis of laboratory values associated with a severe Covid-19 disease progression.

## 2. Methods and patients

### 2.1 Study population and inclusion criteria

For model development we conducted an observational retrospective cohort study using electronic health record data from a hospital of medium level of care located in the federal state of Rhineland-Palatinate in the west of Germany initially collected for billing purposes (Table 1, Figure 1). We included 520 patients with a positive RT-PCR for SARS-CoV-2 identified by the ICD code U07.1 admitted from March 2020 until December 2021 to the hospital. Because of too many missing values, 12 patients were excluded. The missing rate for each feature can be found in the Github repository https://github.com/philippwendland/ML_Covid19. No patient was transferred from an ICU of another hospital. Extreme data points were manually checked for outliers via visual checks using violinplots. No data were removed, because the close inspection revealed that these extreme values are actual measurements.

**Table 1.** Characteristics of the (complete) cohort

| Characteristics | Number (%) or (Q1-Q3) |
| --- | --- |
| Study population | 532 using only 520 |
| In-hospital mortality | 87 (0.167=87/520) |
| Transfer to intensive care unit (ICU) | 89 (0.171) |
| Mechanical ventilation | 59 (0.113) |
| Age | 60.4 (45-82) |
| Female | 248 (0.477) |

**Q1: first quartile, Q3: third quartile**

**Figure 1.**
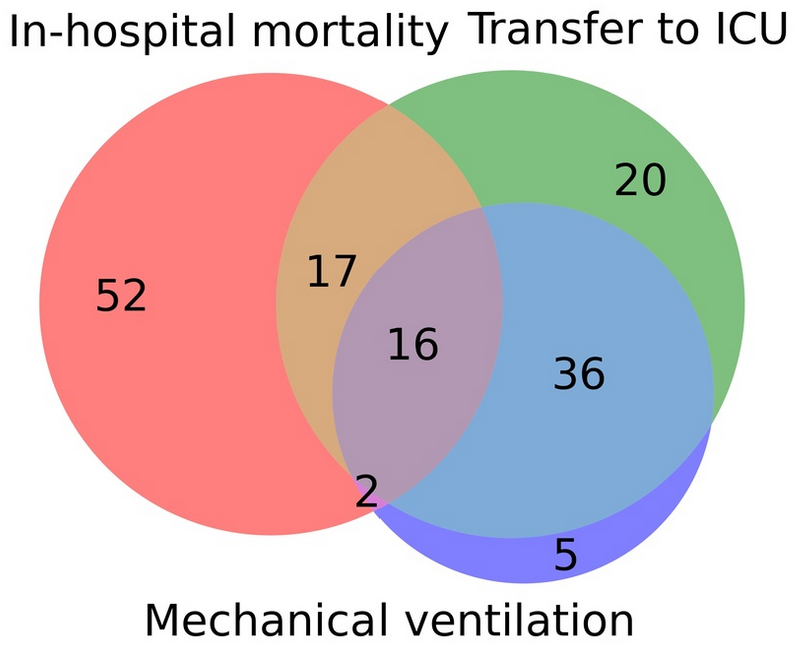
Venn diagram indicating the overlap of the three endpoints “In-hospital mortality”, “transfer to ICU” and “mechanical ventilation” in our cohort from a hospital of medium level of care located in the federal state of Rhineland-Palatinate in the west of Germany.

For model development and prior to any preprocessing steps we performed a random train-test split using 80% of the data as training set and 20% as test set. The report is based on the STROBE-statement, the IJMEDI checklist for assesment of medical AI and the MINIMAR statement [15–17]. Ethical approval was obtained from the local ethics commission of the University of Applied Sciences Koblenz in their 24^th^ meeting.

### 2.2 Study design and statistical analysis

We defined three Covid-19 associated endpoints (see Supplemental Material for details):

1. Death during hospital stay, short “in-hospital mortality”
2. Admission to intensive care unit (ICU), short “transfer to the ICU”
3. Necessity for mechanical ventilation (all OPS beginning with “8-71”), short “mechanical ventilation”

For the training of the prediction models we used laboratory values (see Supplemental Material for a complete list) obtained during the first 48 hours after admission and averaged them over this time period. For prediction and model testing, we restricted the time span to 24 hours after admission. For each endpoint we divided the patient cohort into two distinct groups, depending on whether the endpoint occurred or not. To check for differences in the laboratory values between these groups we performed Wilcoxon-rank-sum tests with Bonferroni-Holm adjusted p-values. The p-values were used as a measure of association strength between the laboratory value and the endpoint and enabled us to rank the features. We filtered the top-10 laboratory values with an adjusted p-value smaller than 5% and less than 10% missing values. These were combined with biological sex and age to form potential features for the machine-learning models. The laboratory values used as covariates were measured by a Cobas 6000 device from Roche Diagnostics, with the exception of PTT, which was measured by a Sysmex CS 2500i from Siemens Healthineers. Xserve was used as laboratory software.

We compared three supervised classifiers: Logistic regression (LR), Random forest (RF) and XGBoost. To select predictive features for each of these three model classes we employed 5-fold cross validation. For LR we performed forward-backward selection. For the random forest classifier and the XGBoost classifier we used the mean feature importance as a criterion for feature selection and in addition also trained these tree based classifiers using the same features as identified for the LR models. Further, for RF and XGBoost we performed a hyperparameter optimization on the training set (see Supplemental Material Section 2.7). The model (including selected features) with the highest receiver operator characteristics area under the curve (ROC-AUC) averaged over the cross validation folds from the training data set was selected as the final model for the respective endpoint.

During model creation we observed that the cross validation performance using the features from the first 48 hours is similar to the performance using the same features observed during the first 24 hours only. Therefore we decided to train our models on data from the first 48 hours, but for prediction and testing we restrict to average laboratory values of the first 24 hours. To handle class imbalance during training we tested the Synthetic Minority Oversampling Technique (SMOTE) [18]. SMOTE creates synthetic patient data of the smaller group in an imbalanced problem resulting in balanced groups. SMOTE did not improve model performance on the validation dataset and therefore our final models are not based on SMOTE (see Supplemental Material section 2.8).

In addition to these models based on numerical laboratory values, we trained models using dichotomous features indicating whether a certain laboratory value exceeds or falls below a predefined reference threshold. In these models, age was also replaced by a dichotomous feature indicating whether the patient was older or younger than 60 years at the time of observation. To prevent data leakage we used the thresholds provided by the manufacturers of the respective laboratory measurement device (see Supplemental Material table S1). These models are easier to interpret and might support the need for rapid decision making by physicians in daily clinical practice (see Supplemental Material for details). We excluded blood sugar from the list of possible dichotomous predictors, because reference values depend on the time gap to the last meal before the blood draw. Information about the last meal was not available. By the very nature of dichotomous variables, these variations have a stronger impact on the status of the feature and might induce unnecessary bias, which is not present in the numerical models.

Calibration is very important, in particular for models applied to clinical decision making [19]. In essence, calibration is the agreement between the estimated probability and the observed frequency of events. To assess calibration, we plotted the observed versus the predicted proportion of events for the respective endpoint (calibration curves) and computed the Brier score. In the calibration curves, we used five bins each containing the same number of samples, which was appropriate for our sample size. Further calibration curves with 10 bins and with bins of identical width are provided in the Supplemental material, section 2.6.

## 3. Results

### 3.1 Study population

A total of 520 patients (248 (47.7%) female) admitted to the hospital between March 2020 and December 2021 and diagnosed with SARS-CoV-2 are included in our study (see Table 1). From these, 87 patients (16.7%) deceased and 89 patients (17.1%) were transferred to the ICU during hospital stay. Due to DNR (Do Not Resuscitate)/DNI (Do Not Inturbate) or palliative treatment just a subgroup of the deceased patients were transferred to the ICU. A mechanical ventilation was performed on 59 patients (11.3%). The mean age of our cohort is 60.4 (45.0 – 82.0), which is expected given that age is a well-known risk factor for severe disease progression [20].

### 3.2 Laboratory values associated with the disease course

For each of the three endpoints we divided the patients into two subgroups, depending on whether the endpoint occurred or not. To identify laboratory values indicating differences between the two respective subgroups we used Wilcoxon-rank-sum-tests with Bonferroni-Holm adjustment. We restricted this to the first 48 hours of the hospital stay and used the adjusted p-values to rank the laboratory values according to their association with the respective endpoint, see Fig. 2. All laboratory values with a p-value smaller than 0.05 were considered to be strongly associated with the endpoint.

**Figure 2.**
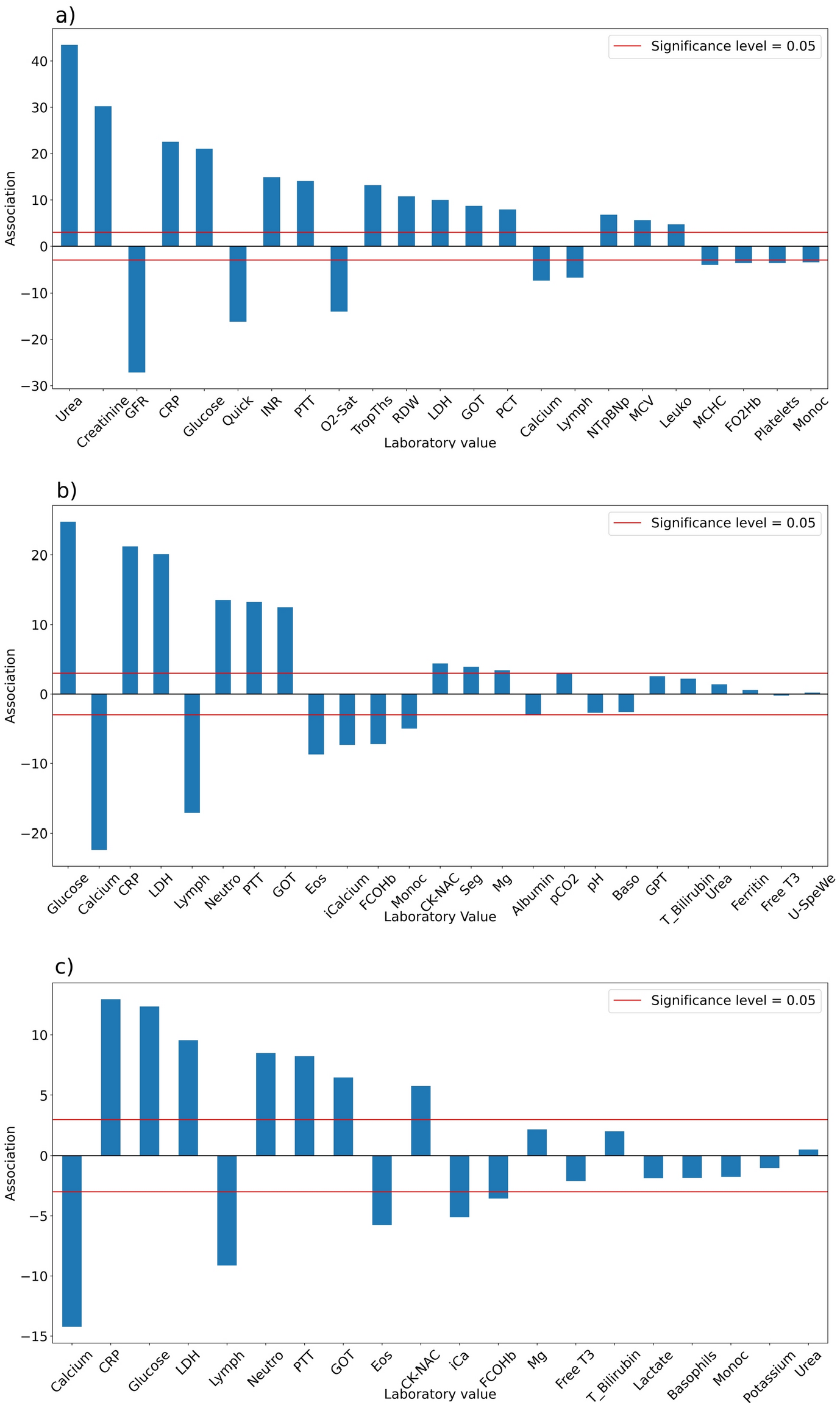
The association between laboratory values and the occurrence of the endpoints a) in-hospital mortality, b) transfer to intensive care unit (ICU) and c) necessity for mechanical ventilation. The association is given by log p-values multiplied by the sign of the association. Positive (negative) values indicate that higher (lower) values of the laboratory values are associated with the endpoint. The p-values are obtained from Wilcoxon rank sum tests for differences in laboratory values of the first 48 hours after admission to a hospital between the two patient groups (Bonferroni-Holm correction for multiple testing). A list of the abbreviations can be found in the Supplemental Material.

For the endpoint in-hospital mortality we found 23 laboratory values to be strongly associated (Fig. 2a). This includes well-known biomarkers for a severe Covid-19 progression, e.g. lymphocytes in % (Lymph) and monocytes in % (Monoc) as hematological biomarkers, CRP in mg/dl, lactatdehydrogenase in mg/dl (LDH) and procalcitonin in ng/ml (PCT) as inflammatory biomarkers and N-terminal of the prohormona brain natriuretic peptide in pg/ml (NTpBNp), glutamic oxaloacetic transaminase in u/l (GOT) as cardiac biomarkers, and calcium in mmol/l as minerals [21]. The laboratory values with the smallest p-values urea in mg/dl and creatinine in mg/dl are known to have elevated levels at admission to hospitals in non-survivors compared to survivors of Covid-19 patients [22]. In accordance with our findings, Covid-19 is sometimes associated with a coagulation dysfunction, which could be indicated through the significant partial thromboplastin time in seconds (PTT), QUICK test in % and INR values [23]. We report significantly increased levels of the mean corpuscular volume in fl (MCV) and decreased levels of the mean corpuscular hemoglobin concentration in g/dl (MCHC) for Covid-19 patients who died during their hospital stay which are known to be altered in Covid-19 patients [24].

For the endpoint transfer to ICU we identified 15 laboratory values (Fig. 2b), nine of them overlapping with the strongly associated laboratory values for in-hospital mortality, including blood sugar in mg/dl (Glucose), calcium and CRP. Interestingly, the two laboratory values urea and creatinine with the smallest p-values for the endpoint in-hospital mortality are not strongly associated with a transfer to the ICU. We identified Neutrophil granulocytes in % (Neutro) to be higher for patients referred to the ICU, but not for patients who died in the hospital. Neutrophil granulocytes were previously reported to play an important role in Covid-19-associated thrombosis [25,26]. Reduced levels of Eosinophils in % (Eos) and an increase in segmented neutrophils in % (Seg) are also strongly associated with a transfer to the ICU, but not with in-hospital mortality. Low ionized Calcium in mmol/l (iCalcium) and calcium are known indicators of a severe Covid-19 disease progression [27].

We found 12 laboratory values to be strongly associated with the necessity for “Mechanical Ventilation” (Fig. 2c). All of them are a subset of the laboratory values strongly associated to transfer to ICU, which makes sense, because most of the patients, who received mechanical ventilation were transferred to the ICU – just seven of them were not transferred to the ICU.

Overall, it can be seen that just a fraction of the 85 to 90 tested laboratory values show a strong association with the endpoints in our population. In agreement with previous reports we find CRP, blood sugar (Glucose), LDH, and Lymph as markers for the occurrence of either of the adverse events. However, it is interesting that urea and creatinine are the laboratory values with the strongest associations to in-hospital mortality, but are not strongly associated with the other two endpoints.

### 3.3 CRP and blood sugar are good predictors for the Covid-19 associated endpoints in-hospital mortality, transfer to the ICU and mechanical ventilation (Figure 3)

We devised prediction models for the occurrence of the endpoints based on biological sex, age and the top-10 laboratory values with the strongest associations to the respective endpoints from Figure 2. We performed 5-fold cross validation on the training data (80%) to select the models and their respective features with the highest ROC-AUC. In Fig. 3 we present results (ROC-curves) for predictions of these selected best models on the test data (20%) not used for training with violinplots of the predictors based on the entire dataset. In-hospital mortality can be predicted from the combination of the three laboratory values CRP, urea and blood sugar evaluated at the first 24 hours after admission augmented by age [20] with an AUC of 0.918 (95% CI: 0.857-0.979) using a logistic regression model (Fig. 3a). Urea as the top laboratory value associated with in-hospital mortality (Fig. 2a) was chosen as a predictive feature, although it is not strongly associated with the other endpoints (Figs 2 b,c).

**Figure 3.**
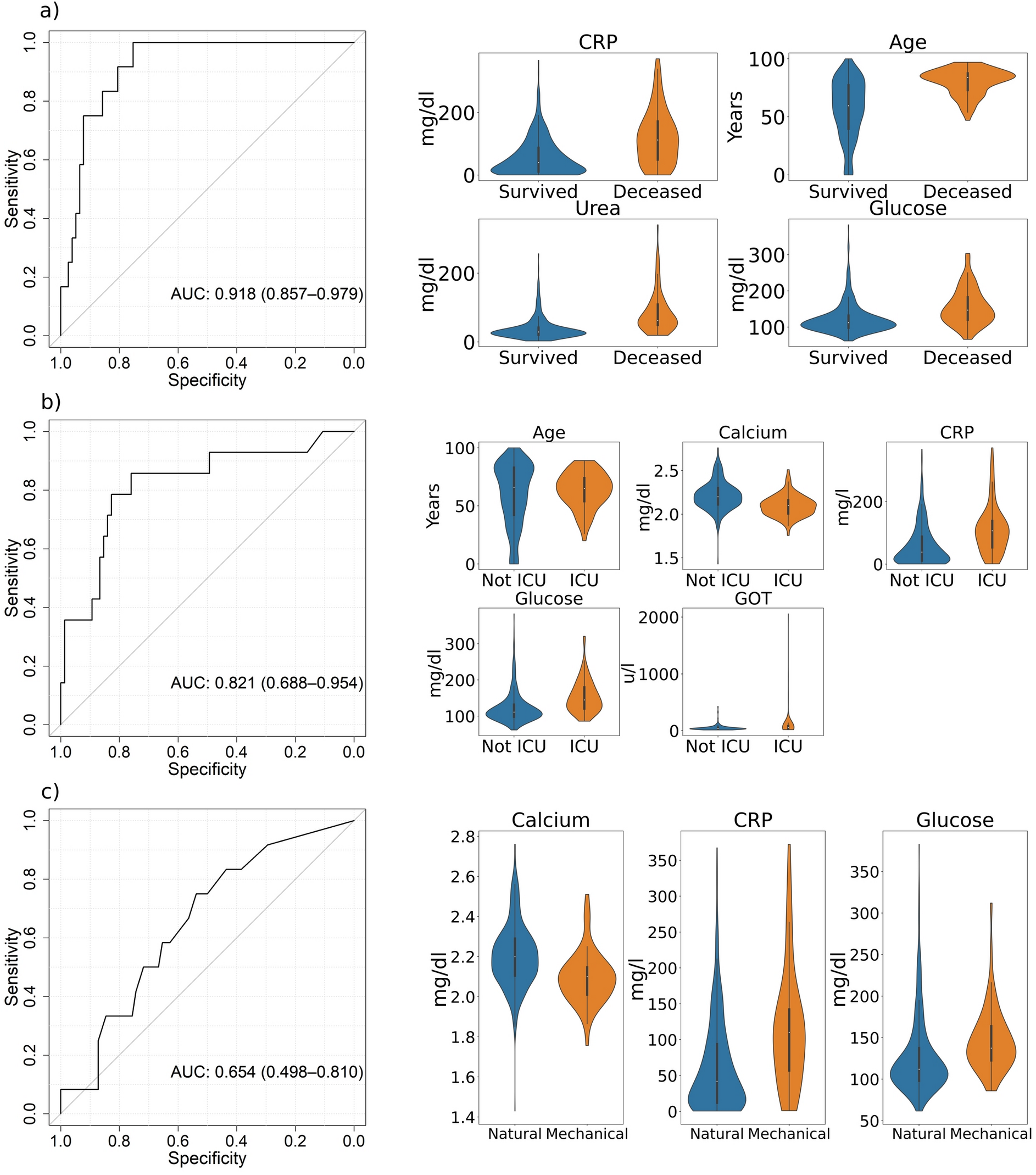
ROC-curves (specificity and sensitivity) of the best predictive machine-learning models for the endpoints **a)** in-hospital mortality (Logistic Regression), **b)** transfer to the ICU (XGBoost), and **c)** mechanical ventilation (Random Forest) with violinplots of their related predictors in the two patient groups. The ROC-curves are based on the test data and the violinplots on the entire dataset. A list of abbreviations can be found in the Supplemental Material.

A more complex nonlinear XGBoost model based on age and four laboratory values predicts the transfer to the ICU (Fig. 3b) with an AUC of 0.821 (95% CI: 0.688-0.954). Please note the differences in the age distribution for this endpoint by contrast with the deceased patients in Fig. 3a. Compared to this endpoint, the laboratory values GOT and calcium were chosen in addition to CRP and blood sugar as predictors for a transfer to the ICU, whereas urea was eliminated by the feature selection procedure. Some patients exhibit extreme GOT levels, as indicated by the violin plots.

Most patients who were transferred to the ICU also received mechanical ventilation. Nevertheless, prediction of mechanical ventilation is more difficult (Fig. 3c). The best model is a Random Forest based on calcium, CRP and blood sugar with a test AUC of 0.654 (95% CI: 0.498-0.81). These laboratory values are also in the set of predictors for a transfer to the ICU. Increased levels of CRP and blood sugar are strongly associated with and important predictors for all three endpoints.

### 3.4 PTT and GOT are good dichotomous predictors for the Covid-19 associated endpoints in-hospital mortality, transfer to the ICU and mechanical ventilation (Figure 4)

The combination of numerical laboratory values and age might still not be simple enough to guide medical decision making under stressful conditions in hospitals. Therefore, we devised models based on dichotomous features indicating, whether the value is higher or lower than a predefined critical threshold. In addition, we also used a dichotomous feature for age, indicating whether the patient was younger than 60 years or not.

**Figure 4.**
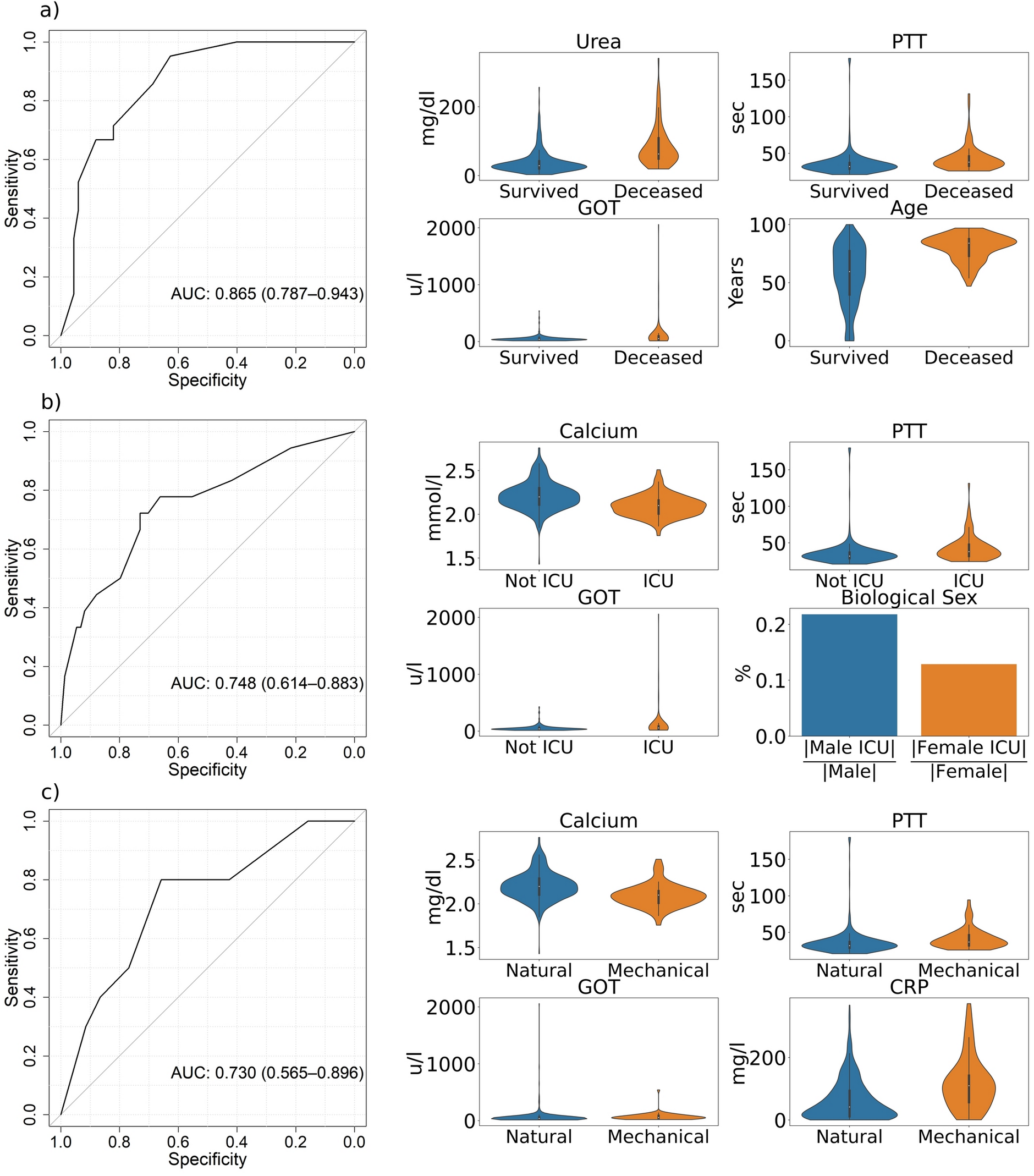
ROC-curves (specificity and sensitivity) of the best predictive machine-learning models based on dichotomous predictors regarding the endpoints **a)** in-hospital mortality (Logistic Regression), **b)** transfer to the ICU (Logistic Regression), and **c)** mechanical ventilation (XGBoost) with violinplots of their respective predictors. Biological sex describes the fraction of all male/female patients with Covid-19, who were transferred to the ICU. The ROC-curves are based on the test data and the violinplots on the entire dataset. A list of abbreviations can be found in the Supplemental Material.

In hospital mortality can be predicted from dichotomous values for urea, PTT, GOT and age by logistic regression with an AUC of 0.865 (95% CI: 0.787-0.943), see Fig. 4a. This is only slightly worse than the prediction from numerical features (compare Fig. 3a). Age and urea are included as predictors in both the numerical and dichotomous model for this endpoint.

Using only dichotomised features, transfer to the ICU can be predicted with an average AUC of 0.748 (95% CI: 0.614 to 0.883), see Fig. 4b. This is nearly as accurate as the prediction from numerical features (compare Fig. 3b). The selected logistic regression model uses the laboratory values calcium, PTT and GOT in combination with biological sex as predictors (Fig 4b). GOT and calcium are also part of the numerical model.

Predicting the necessity of mechanical ventilation using dichotomous features only (Fig. 4c) seems to be not less accurate (AUC of 0.73, 95% CI:0.565-0.896) than predictions from numerical features (Fig. 3b). For this endpoint, the best performing model is again XGBoost with calcium, CRP, PTT and GOT as predictors. Calcium and CRP was also selected in the model with numerical features, whereas blood sugar was replaced by a combination of PTT and GOT in the model with dichotomous features only. As for the numerical features, neither age nor biological sex as additional features improved the prediction (cross validation on the training data) of the need for mechanical ventilation.

The differences between the features selected for the numerical and dichotomous models indicate that some laboratory values are more suitable for decisions based on dichotomized values (“too high / too low”) than others. The reference range of the laboratory values is defined such that 95% of a healthy reference population have values lying within the reference range, which does not mean, that laboratory values lying outside the reference range are automatically critical values [28]. For example, urea and calcium seem to be robust against dichotomization, whereas the absolute level of the CRP seems to be more informative than just an increase above the reference level. In contrast, a too high a value of PTT seems to be informative even when the absolute level is not considered.

### 3.5 Calibration and additional performance metrics

To assess model calibration we provide calibration curves of the prediction models (see Figure 5), which compare the probability of a given class predicted by the model to the observed fraction. Ideally, these are equal and the calibration curves are diagonal.

**Figure 5.**
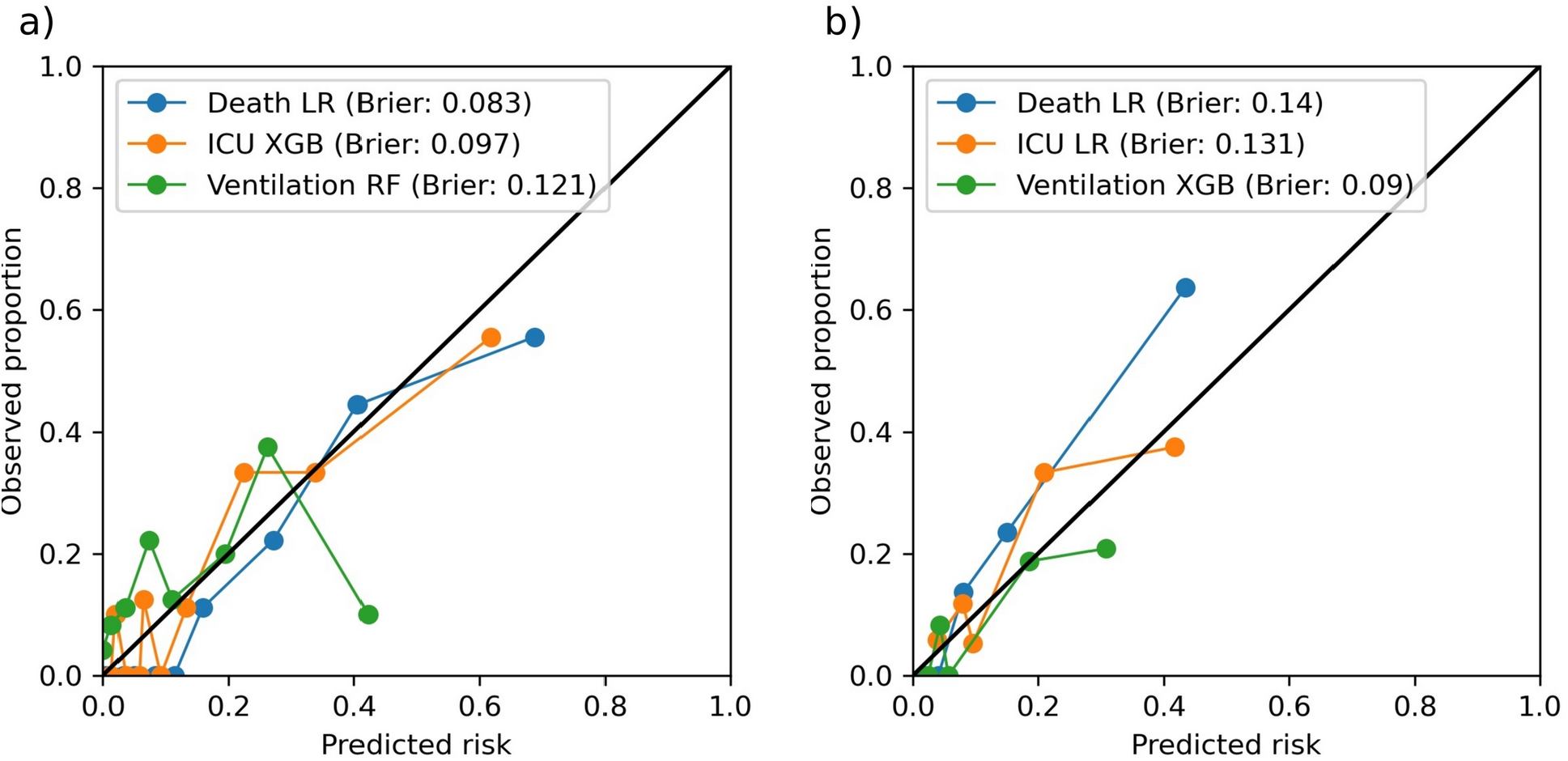
Calibration curves including Brier scores of the best predictive machine-learning models **a)** based on numerical predictors and **b)** based on dichotomous predictors for the endpoints in-hospital mortality, transfer to the ICU and mechanical ventilation.

Deviations from the diagonal were quantified by the Brier score, which is the mean squared error relative to the diagonal.

These calibration curves were obtained for the test data after training the models.

The prediction model for in-hospital mortality based on numerical covariates has a Brier score of 0.083, but it seems that the model slightly overestimates risks. In contrast, the prediction model for in-hospital mortality based on dichotomous covariates has a Brier score of 0.14 and slightly underestimates risks. The model for predicting ICU admission based on numerical features is well calibrated (Brier score = 0.091), whereas the model based on dichotomous features slightly underestimates risks. Both models for predicting mechanical ventilation overestimate the risk, which is possibly caused by the small sample size. Only 59 patients in our cohort were mechanically ventilated (Figure 1) and the calibration curves might not be very reliable estimates.

Overall, the models for the endpoints in-hospital mortality and ICU admission are reasonably well calibrated, which can also be seen from the expected calibration error [29] of the different models (see Tables 2 and 3) and we abstain from recalibrating the models. For mechanical ventilation, the calibration curves can not be reliably estimated because of the small number of events in the data (see Supplemental material section 2.6 for calibration curves including 10 bins and bins with identical widths). Whilst the ROC curves in Figures 3 and 4 provide information about the sensitivity and specificity trade-off of our prediction models, there are a number of alternative performance metrics which emphasize other aspects [30,31]. In Tables 2 and 3 we report the negative and positive predictive values, the F1 score, the accuracy and the balanced accuracy. These precision metrics depend on the specific threshold chosen for the score (or the estimated class conditional probability) of the prediction model. In Table 2, we chose a threshold of 50% as a decision rule for the two classes. In Table 3 we defined the binary decision boundary by optimizing the F-1 score for the training set. The results in Table 3 indicate, that for an optimal trade-off between precision and recall we still observe a relatively good balanced accuracy, despite the class imbalance in our training and test set. Please note, that the positive and negative predictive value depend on the prevalence of the respective endpoints and might change in a different cohort. However, for our cohort, the high values of the negative predictive values indicate that we can safely identify the low risk patients.

**Table 2.** Additional performance metrics for the different prediction models estmated for the test data set. The decision boundary was defined by a threshold of 50% for the estimated class conditional probability. Abbreviations: AUC: ROC-AUC (see Figures 3 for numerical and 4 for dichotomous features), Brier score for calibration, ECE for expected calibration error, NPV and PPV for negative and positive predictive value, F-1 for F-1-score, ACC for accuracy and BalAcc for balance accuracy.

| Model | AUC | Brier | ECE | NPV | PPV | F-1 | ACC | Bal Acc |
| --- | --- | --- | --- | --- | --- | --- | --- | --- |
| Death | 0.918 | 0.083 | 0.054 | 0.901 | 0.5 | 0.4 | 0.865 | 0.64 |
| ICU | 0.821 | 0.097 | 0.041 | 0.881 | 0.8 | 0.421 | 0.876 | 0.636 |
| Ventilation | 0.654 | 0.121 | 0.088 | 0.874 | 0.333 | 0.13 | 0.855 | 0.523 |
| Death (dichot.) | 0.865 | 0.14 | 0.096 | 0.78 | 0.5 | 0.222 | 0.761 | 0.549 |
| ICU (dichot.) | 0.748 | 0.131 | 0.015 | 0.83 | 0.75 | 0.273 | 0.826 | 0.577 |
| Ventilation (dichot.) | 0.73 | 0.089 | 0.028 | 0.914 | 0.3 | 0.3 | 0.848 | 0.607 |

**Table 3.** The same performance metrics as in Table 2 for the test data, but here the decision boundary was defined by a maximum F-1 score, determined for the training set.

| Model | AUC | Brier | ECE | NPV | PPV | F-1 | ACC | Bal Acc |
| --- | --- | --- | --- | --- | --- | --- | --- | --- |
| Death | 0.918 | 0.083 | 0.054 | 0.96 | 0.6 | 0.666 | 0.898 | 0.836 |
| ICU | 0.821 | 0.097 | 0.041 | 0.954 | 0.458 | 0.579 | 0.82 | 0.806 |
| Ventilation | 0.654 | 0.121 | 0.088 | 0.90 | 0.214 | 0.3 | 0.689 | 0.609 |
| Death Cat (dichot.) | 0.865 | 0.14 | 0.096 | 0.894 | 0.636 | 0.651 | 0.83 | 0.774 |
| ICU Cat (dichot.) | 0.748 | 0.131 | 0.015 | 0.916 | 0.394 | 0.433 | 0.728 | 0.726 |
| Ventilation (dichot.) | 0.73 | 0.089 | 0.028 | 0.964 | 0.222 | 0.348 | 0.674 | 0.729 |

## 4. Discussion

### 4.1 Comparison to other studies

There are several data driven models for the prediction of severe COVID-19 disease courses with different setting and goals. Here, we compare our results to some of the previous approaches.

Famiglini et al. [32] devised several machine-learning models predicting ICU admission of COVID-19 patients within the next five days using gender, age and the complete blood count as potential features. Their best model achieves a ROC-AUC of 0.85 and a Brier score of 0.144. Our numerical model predicts ICU submission from data of the first 24 hours after admission and provides similar accuracy with only a few predictors and is also well calibrated. Our predictors were selected from a large set of laboratory values in an unbiased way. In addition, we have models using only dichotomous predictors which are very easy to interpret.

Campbell et al. [33] devised hierarchical ensemble classification models for the prediction of several severe events connected with Covid-19 based on laboratory and clinical data available at admission. Due to missing values they removed about 50 percent of the training data and 85 percent of test data. This might be the reason for the limited performance of these models on test data.

Wu et al. [8] created a logistic regression model for predicting “severe disease” of hospitalized Covid-19 patients with an extensive external validation on five datasets with a mean ROC-AUC of 0.88, mean sensitivity of 0.85 and mean specificity of 0.74. Patients were labeled as “severely diseased” if they die, get a shock, were admitted to the ICU, develop organic failure or were mechanically ventilated during hospital stay. Their model is based on demographic features, symptoms, laboratory values and radiological findings.

We have developed models based on standard laboratory values, age and biological sex for more specific endpoints using only data from the first 24 hours after hospital admission. In addition, we think that the dichotomous models are an useful addition to these studies. Wollenstein-Betech et al. [7] devised machine-learning prediction models for adverse events of Covid-19 patients using publicly available data of 91.000 patients from Mexico.

Their models were based on demographic features and comorbidities leading to a ROC-AUC of 0.75 for hospitalization and 0.7 for mortality. Although their work is different in scope, it is remarkable that simple models like logistic regression and support vector machines perform just as well as more complex models. They used both positive Covid-19 patients and patients waiting for test results, which might induce label noise. In our data, we could see a clear difference in the distribution of many laboratory values in patients with a positive PCR test and patients that were only diagnosed with Covid-19 based on the clinical impression, but did not have a positive test.

Heber et al. [14] developed a linear mixed model for the prediction of in-hospital mortality using patient-specific intercepts and slopes of hematological parameters measured during the first 4 days after admission. With a ROC-AUC of 0.92 their model achieves similar performance like ours, but our model is tested on laboratory values of the first 24 hours after admission. Further, we prevent bias by using a data-driven feature selection, whereas Heber et al. limit their set to just twelve potential variables before feature selection.

Häger et al. [10] report an external validation of the predictive performance for five important Covid-19 risk scores for in-hospital mortality and ICU admission. The 4c-score [34] performs best for in-hospital mortality with a ROC-AUC of 0.81 and the easy-to-use bedside score NEWS [35] performs best for ICU admission with a ROC-AUC of 0.83. Our data-driven models perform better for in-hospital-mortality and similarly for ICU admission. Son et al. [3] provides a simple and well interpretable four class score for predicting severity of Covid-19 based on vital parameters, but unfortunately do not test the performance on patient data.

Recent reviews [36,37] describe the landscape of state-of-the-art machine-learning models applied to Covid-19 patient data. Most papers use quite simple models based on Logistic Regression, XGBoost and Support Vector machines. As pointed out in [31], a limitation of many studies is that the data base for these models includes sicker patients, which could potentially result in selection bias.

In comparison to previous work, our models are based on an unbiased set of potential features and result in simple models. Another new contribution are the models with dichotomous features, which are easy to apply and to interpret, without substantially impeding the predictive performance.

### 4.2 Summary and Conclusion

All in all, we devise purely data-driven predictive machine-learning models for a severe Covid-19 outcome using a small and well interpretable number of standard laboratory values combined with age and biological sex. The endpoints in-hospital mortality and transfer to the ICU can be predicted with high or good accuracy within the first 24 hours after admission. Predicting the need for mechanical ventilation is much more difficult. For all three endpoints, models using only dichotomous features perform only slightly worse than models based on a complex combination of numerical laboratory values, sometimes complemented by age and/or biological sex. In particular, the models based on dichotomous features are simple to interpret and easily applicable in a real life hospital setting. Further, the simplicity of our models offers a real-time online prediction of the patient risks with prediction times far less than one second.

For some laboratory values including CRP and blood sugar the numerical values are informative for prediction, whereas other laboratory values like PTT and GOT are suitable as dichotomous features indicating values which are too high or too low. We observe that many features including CRP, blood sugar, LDH and Lymph are strongly associated to all of the three endpoints. Intriguingly, urea and creatinine are the laboratory values most strongly associated with in-hospital mortality, although they are not significantly associated with the other two endpoints.

A real world application of our models as a risk assessment tool requires to define thresholds with a reasonable trade off between sensitivity and specificity. Depending on the real but unknown prevalence of the high risk patient group, the positive and negative predictive value at a given threshold might change, when our tests are applied in a different hospital or in a different phase of the pandemic. The high intrinsic sensitivity of our model for detecting patients at risk of death (Figure 3 a) with a moderate specificity implies that the negative predictive value is still high, enabling us to safely stratify patients with low risk (compare also Tables 2 and 3). For high risk patients, a close monitoring and possibly, depending on further diagnostics and possible drug interactions, a treatment with Remdesivir or Nirmatrelvir/Ritonavir and a prophylactic dose of heparin might be considered.

The features included in the dichotomous models might also be useful on its own. For a patient who has a risky pattern of these dichotomous features, the laboratory software or the laboratory technicians can already assign a warning and make doctors aware that this patient might need closer monitoring. Potentially, this could support the decision making for antibody or antiviral treatment, in combination with other diagnostic results of the individual patient.

We also analyzed ICD codes for diagnosis as additional features and found significant differences between the two patient groups for each endpoint (Supplemental Material Figure S2). However, the inclusion of these diagnostic features did not improve the models much. This suggests that laboratory values alone are sufficient to predict Covid-19 outcomes in hospitals. In addition, the time of diagnosis is often not available in our data.

### 4.3 Limitations

In our study we include patients admitted to hospitals from the beginning of the pandemic until the end of 2021. Due to the rapidly changing epidemiological circumstances of the pandemic we were not able to test the generalizability of our models to a population, where the Omicron mutation is the dominating virus mutation. From 2020 until December 2021 the Wildtype, Alpha, Beta and Delta mutations were the dominating Covid-19 variants in Germany [38,39]. Unfortunately, we have no opportunity to check the patient-level mutation status of the virus variant, but it is plausible that these might be the dominating mutations in our dataset.

Furthermore, we have no data regarding the vaccination status of the patients, but we assume that most patients until spring or summer 2021 were not completely vaccinated against Covid-19, but after summer 2021 the majority of the patients should be completely vaccinated based on the vaccination rate in Germany [40].

The inclusion of vital parameters, pre-existing comorbidites and vaccination status could improve our models. Unfortunately, due to missing information we are not able to remove patients with DNR/DNI, which could induce a bias in our prediction models.

The cohort of 520 patients is relatively small. The uncertainty of our ROC-AUCs can be reduced by a larger sample size. Further, our data is imbalanced, because only 50 to 90 patients with poor outcomes were observed for each respective endpoint. Although we adressed this issue, we can not completely exclude bias induced by this class imbalance.

### 4.4 Outlook

To test how well our predictions generalize to other hospitals, we will evaluate the performance of the trained models on a test set from a different patient cohort and different hospitals. This will also include extensions to patient cohorts with other dominating virus mutations. Further improvements include time dependent predictions allowing for an online monitoring of patients, taking the patient history into account. We will also check whether the incorporation of genetic risk factors associated with a severe Covid-19 progression [41] can improve the predictions even further.

## Data Availability

Raw data can not made available because of German and EU data privacy regulations.

## 4.7 Ethical Approvement Statement

The restrospective observational study was approved by and performed according to the guidelines of the local ethics committees.

## 4.8 Data Availability Statement

Due to german data protection law we are not allowed to publicly share the patient data used in this publication.

