## Supplemental Information for "Machine learning models for predicting severe COVID-19 outcomes in hospitals"

Additional supplemental materials including the code to reproduce the results can be found in our Github repository https://github.com/philippwendland/ML_Covid19.

### 1. Supplemental material regarding Methods

#### *1.1 Abbreviations and units*

*A list of the abbreviations of all tested laboratory values and their corresponding units can be found in our Github repository as "lab_abbrevations.csv".*

#### *1.2 Information about used packages*

All analysis was made in python and R with the following packages pandas [1], numpy [2], matplotlib [3], seaborn [4], scipy [5], statsmodels [6], sklearn [7], scikit-optimize [8], xgboost [9], tidyverse [10], and proc [11].

#### *1.3 Information about the workflow of the study*

To simplify understanding we present a schematic workflow of the study as a flow chart in the supplemental information.

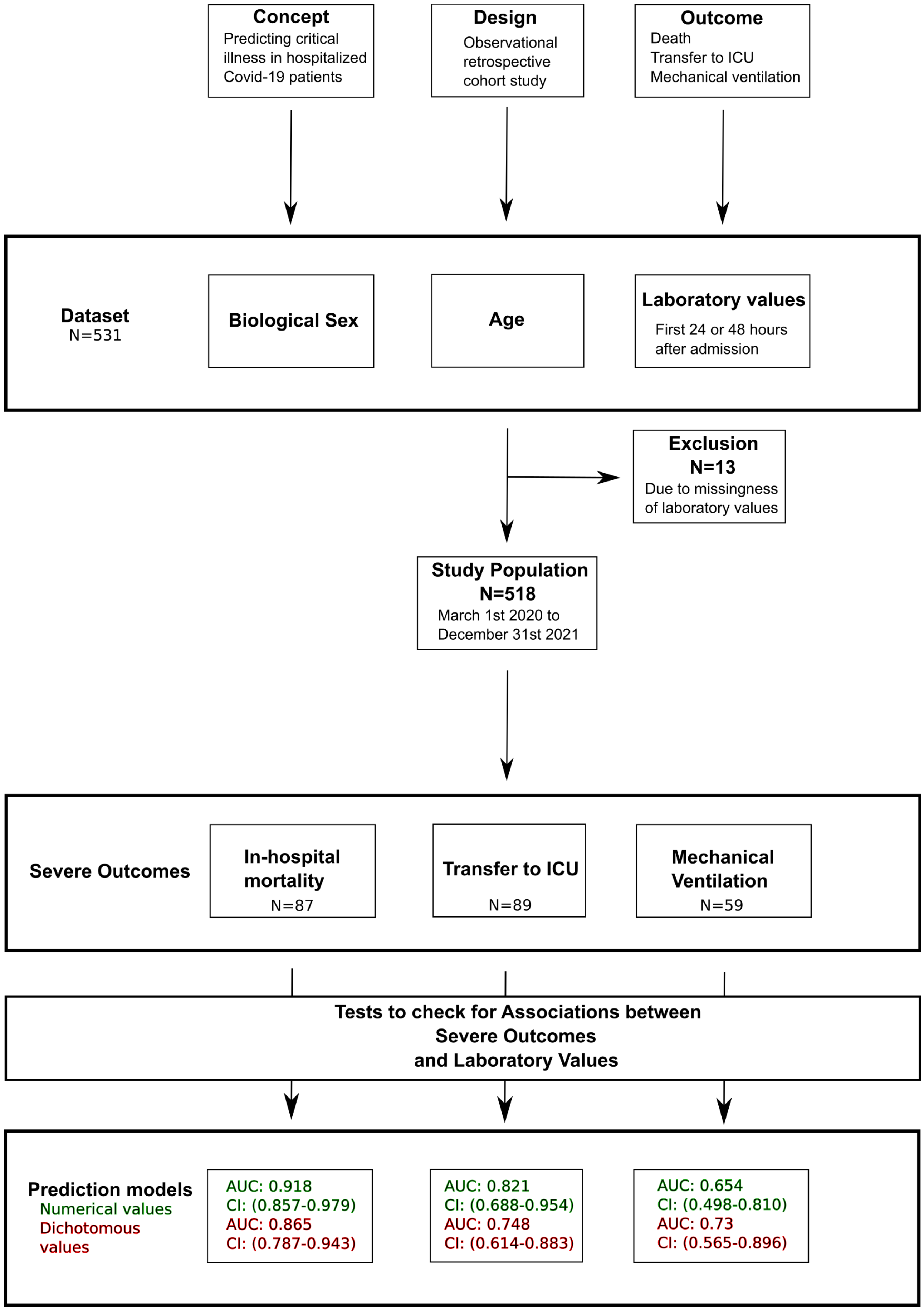


**Figure S1.** Schematic workflow presenting the steps of our study.

*1.4 Information about OPS*

The “Operationenschlüssel nach §301 SGB V” (OPS-301) has been used since 1996 for the documentation of diagnoses and procedures in german hospitals. It is included in the ICPM-GE, which is based on the ICPM Dutch Extension of the original ICPM, which was published by the World Health Organization [12,13].

*1.5 Additional Information about the endpoint mechanical ventilation*

OPS codes starting with ”8-71” are part of the category ”Maschinelle Beatmung und Atemunterstützung über Maske oder Tubus und Beatmungsentwöhnung” (Mechanical ventilation and respiratory support via mask or tube and ventilation weaning).

All the OPS codes regarding mechanical ventilation are linked to patients of intensive care. Patients with mechanical ventilation started during operation with duration less than 24 hours were excluded. Every patient but one got high flow nasal cannula (HFNC) – (the hospital do not offer Extracorporeal membrane oxygenation (ecmo)).

*1.6 Additional Information about the binarized features*

The reference range of the laboratory values is defined such that, for 95% of a healthy reference population the laboratory values are lying within reference range. To create our binarized features we apply t-tests to our data to get the information, whether it is critically if a variable is above the upper reference threshold or underneath the lower reference threshold. To prevent data leakage we use the thresholds provided by the manufacturer of the laboratory measuring instruments. We present the laboratory values including reference ranges based on the covariates age and biological sex with information about sources of the thresholds in Table S1. An empty cell means, that the reference values are similar regarding this covariate.

**Table S1**
Laboratory values including reference ranges based on age and biological sex with information about the critical threshold including sources of the thresholds

| Laboratory value | Biological sex | Age | Lower range | Upper range | Critical Threshold | Sources of the thresholds |
|---|---|---|---|---|---|---|
| Urea | | -1month | 6 mg/dl | 41 mg/dl | Upper | Roche Diagnostics |
| Urea | | 1month-1year | 10 mg/dl | 43 mg/dl | Upper | Roche Diagnostics |
| Urea | | 1year- | 10 mg/dl | 50 mg/dl | Upper | Roche Diagnostics |
| PTT | | -4weeks | 31 Sec | 54 Sec | Upper | Siemens-Healthineers |
| PTT | | 4weeks-3months | 25 Sec | 60 Sec | Upper | Siemens-Healthineers |
| PTT | | 3months-9months | 27 Sec | 50 Sec | Upper | Siemens-Healthineers |
| PTT | | 9months-5years | 24 Sec | 36 Sec | Upper | Siemens-Healthineers |
| PTT | | 5years- | 25 Sec | 38 Sec | Upper | Siemens-Healthineers |
| GOT | F | | 10 u/l | 35 u/l | Upper | Roche Diagnostics |
| GOT | M | | 10 u/l | 50 u/l | Upper | Roche Diagnostics |
| Calcium | | -1month | 1.8 mmol/l | 2.8 mmol/l | Lower | Roche Diagnostics |
| Calcium | | 1month-1year | 2.1 mmol/l | 2.7 mmol/l | Lower | Roche Diagnostics |
| Calcium | | 1year-14years | 2.1 mmol/l | 2.6 mmol/l | Lower | Roche Diagnostics |
| Calcium | | 14years-60years | 2.11 mmol/l | 2.55 mmol/l | Lower | Roche Diagnostics |
| Calcium | | 60years-90years | 2.1 mmol/l | 2.55 mmol/l | Lower | Roche Diagnostics |
| Calcium | | 90years- | 2.05 mmol/l | 2.4 mmol/l | Lower | Roche Diagnostics |
| CRP | | | 0 mg/l | 5 mg/l | Upper | Roche Diagnostics |

### 2 Supplemental material regarding Results

#### *2.1 Example of all covariate combinations for binary logistic regression model*

Using n binary covariates lead to $2^n$ possible outcomes of our predictive models. As an example we show all possible binary covariate combinations and their related outcomes of our logistic regression model for the prediction of in-hospital mortality (see Table S1). Via

Youden’s Index it is possible to determine an optimal threshold of a classifier [14]. With an threshold  and the table of all possible outcomes it is possible to detect rapidly and easily patients with a high risk of in-hospital mortality.

**Table S2**

All possible outcomes of our logistic regression model for the prediction of in-hospital mortality based on the binarized variables of Urea, PTT, Age and GOT

| Urea | PTT | Age | GOT | Outcome |
|---|---|---|---|---|
| 0 | 0 | 0 | 0 | 0.3286146434080142 |
| 1 | 0 | 0 | 0 | 0.10498947201797815 |
| 0 | 1 | 0 | 0 | 0.13992264396094614 |
| 0 | 0 | 1 | 0 | 0.6689658284899486 |
| 0 | 0 | 0 | 1 | 0.19014819252697368 |
| 1 | 1 | 0 | 0 | 0.037526813495647945 |
| 1 | 0 | 1 | 0 | 0.3262914665868695 |
| 1 | 0 | 0 | 1 | 0.05327379143127271 |
| 0 | 1 | 1 | 0 | 0.40180167432851727 |
| 0 | 1 | 0 | 1 | 0.07239138116390866 |
| 0 | 0 | 1 | 1 | 0.49223095810753353 |
| 0 | 1 | 1 | 1 | 0.24369011971541504 |
| 1 | 0 | 1 | 1 | 0.1885290372822444 |
| 1 | 1 | 0 | 1 | 0.018360175032780132 |
| 1 | 1 | 1 | 0 | 0.13865794558658076 |
| 1 | 1 | 1 | 1 | 0.07168619305898585 |

#### *2.2 Additional Plots of the associations of the diagnosis*

To check for differences between the frequency of diagnosis we apply Fisher‘s exact test. Although significant differences between the frequency of diagnosis between the two patient groups for each endpoint were observed (Figure S1), we found that inclusion of these diagnostic features did not improve the models much. Due to missing values of time of diagnosis it was not possible to restrict the inclusion for diagnosis data to 24 or 48 hours after admission.

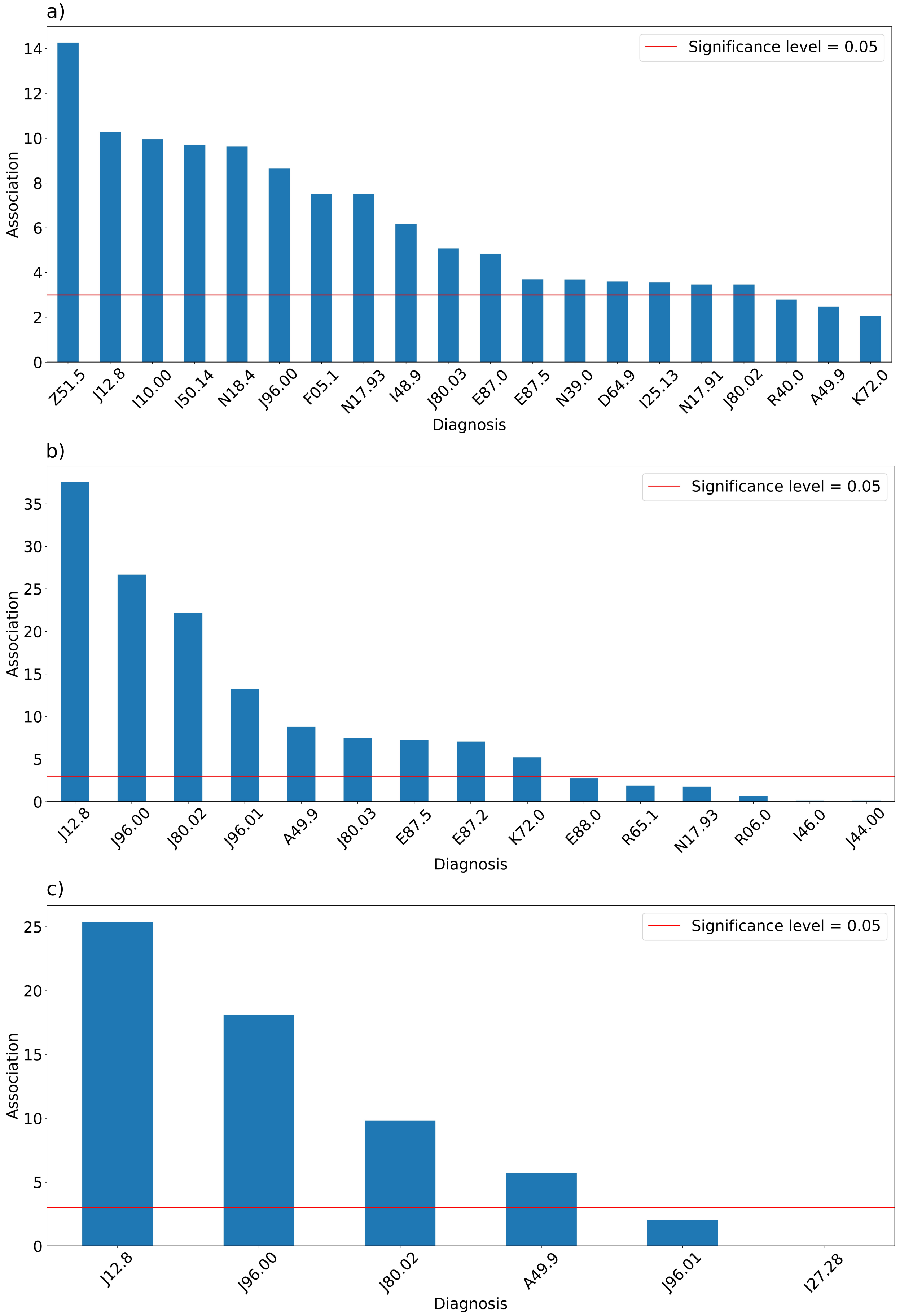
a)
Significance level = 0.05
Association
Diagnosis
Z51.5
J12.8
I10.00
I50.14
N18.4
J96.00
F05.1
N17.93
I48.9
J80.03
E87.0
E87.5
N39.0
D64.9
I25.13
N17.91
J80.02
R40.0
A49.9
K72.0
b)
Significance level = 0.05
Association
Diagnosis
J12.8
J96.00
J80.02
J96.01
A49.9
J80.03
E87.5
E87.2
K72.0
E88.0
R65.1
N17.93
R06.0
I46.0
J44.00
c)
Significance level = 0.05
Association
Diagnosis
J12.8
J96.00
J80.02
A49.9
J96.01
I27.28

**Figure S2.** The association between diagnosis as ICD-10 codes and the occurrence of the endpoints a) in-hospital mortality, b) transfer to intensive care unit (ICU) and c) necessity for mechanical ventilation. The association is given by p-values obtained from fisher's exact test (Bonferroni-Holm correction for multiple testing).

#### *2.3 Wilcoxon-rank-sum-tests and t-tests*

Results of all tests regarding the endpoints can be found in our Github Repository under the name "wilcoxon_covid_dead48h.csv", "wilcoxon_covid_icu48h.csv", "wilcoxon_covid_vent48h.csv", "ttest_covid_dead48h.csv", "ttest_covid_icu48h.csv" and "ttest_covid_vent48h.csv".

#### *2.4 Additional Plots of the associations of the laboratoy values*

In addition to the barplots of the logarithm of the Bonferroni-Holm adjusted p-values we present plots of the not-logarithmized Bonferroni-Holm adjusted p-values.

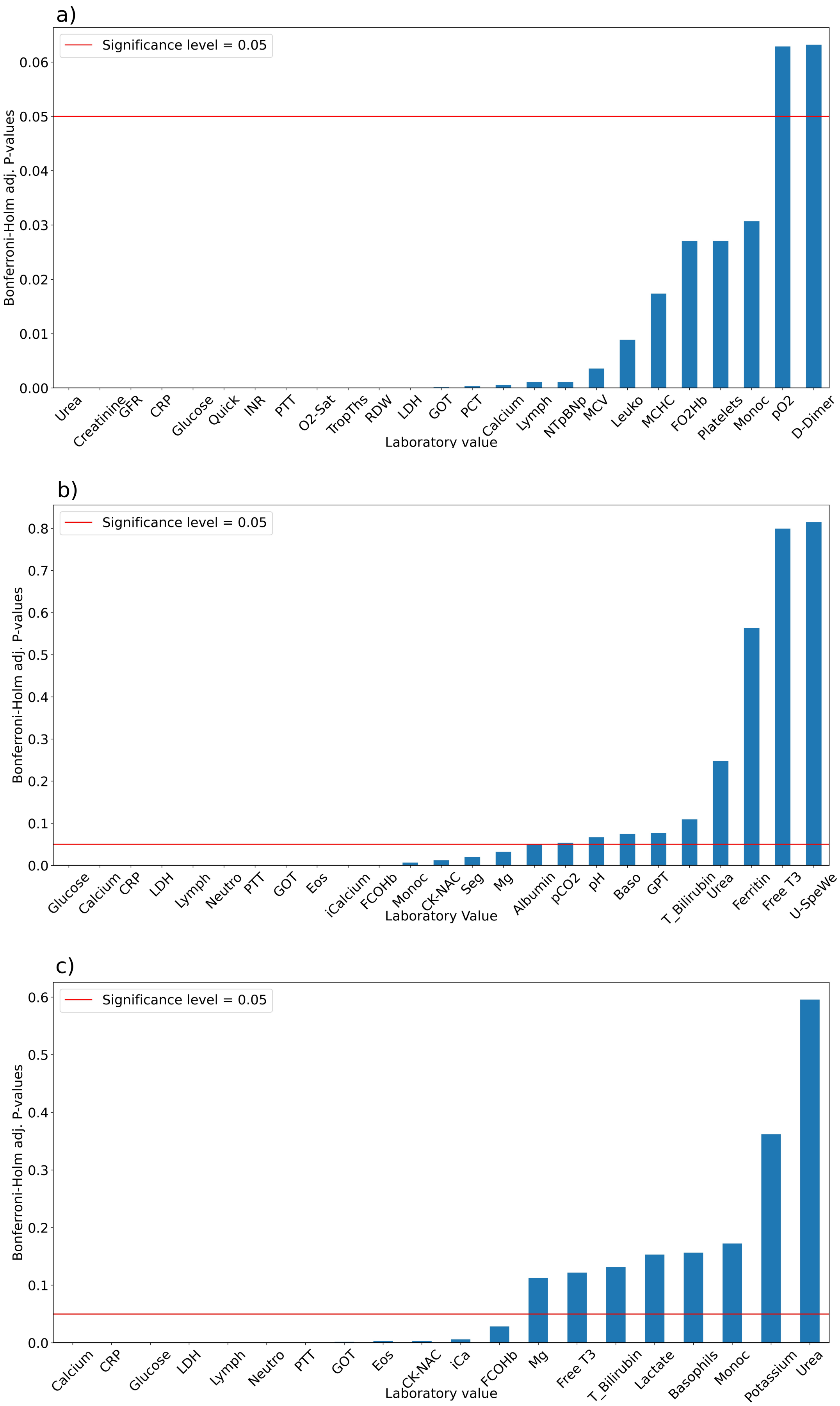
a)
Significance level = 0.05
Bonferroni-Holm adj. P-values
0.06
0.05
0.04
0.03
0.02
0.01
0.00
Urea
Creatinine
GFR
CRP
Glucose
Quick
INR
PTT
O2-Sat
TropThs
RDW
LDH
GOT
PCT
Calcium
Lymph
NTpBNp
MCV
Leuko
MCHC
FO2Hb
Platelets
Monoc
pO2
D-Dimer
Laboratory value
b)
Significance level = 0.05
Bonferroni-Holm adj. P-values
0.8
0.7
0.6
0.5
0.4
0.3
0.2
0.1
0.0
Glucose
Calcium
CRP
LDH
Lymph
Neutro
PTT
GOT
Eos
iCalcium
FCOHb
Monoc
CK-NAC
Seg
Mg
Albumin
pCO2
pH
Baso
GPT
T_Bilirubin
Urea
Ferritin
Free T3
U-SpeWe
Laboratory Value
c)
Significance level = 0.05
Bonferroni-Holm adj. P-values
0.6
0.5
0.4
0.3
0.2
0.1
0.0
Calcium
CRP
Glucose
LDH
Lymph
Neutro
PTT
GOT
Eos
CK-NAC
iCa
FCOHb
Mg
Free T3
T_Bilirubin
Lactate
Basophils
Monoc
Potassium
Urea
Laboratory value

**Figure S3.** The association between laboratory values and the occurrence of the endpoints a) in-hospital mortality, b) transfer to intensive care unit (ICU) and c) necessity for mechanical ventilation. The association is given by p-values obtained from Wilcoxon rank sum tests for differences in laboratory values of the first 48 hours after admission to a hospital between the two patient groups (Bonferroni-Holm correction for multiple testing).

#### *2.6 Additional calibration curves*

We present additional calibration curves using ten bins each having the same number of patients and using five bins uniformly spaced (see Figure S4).

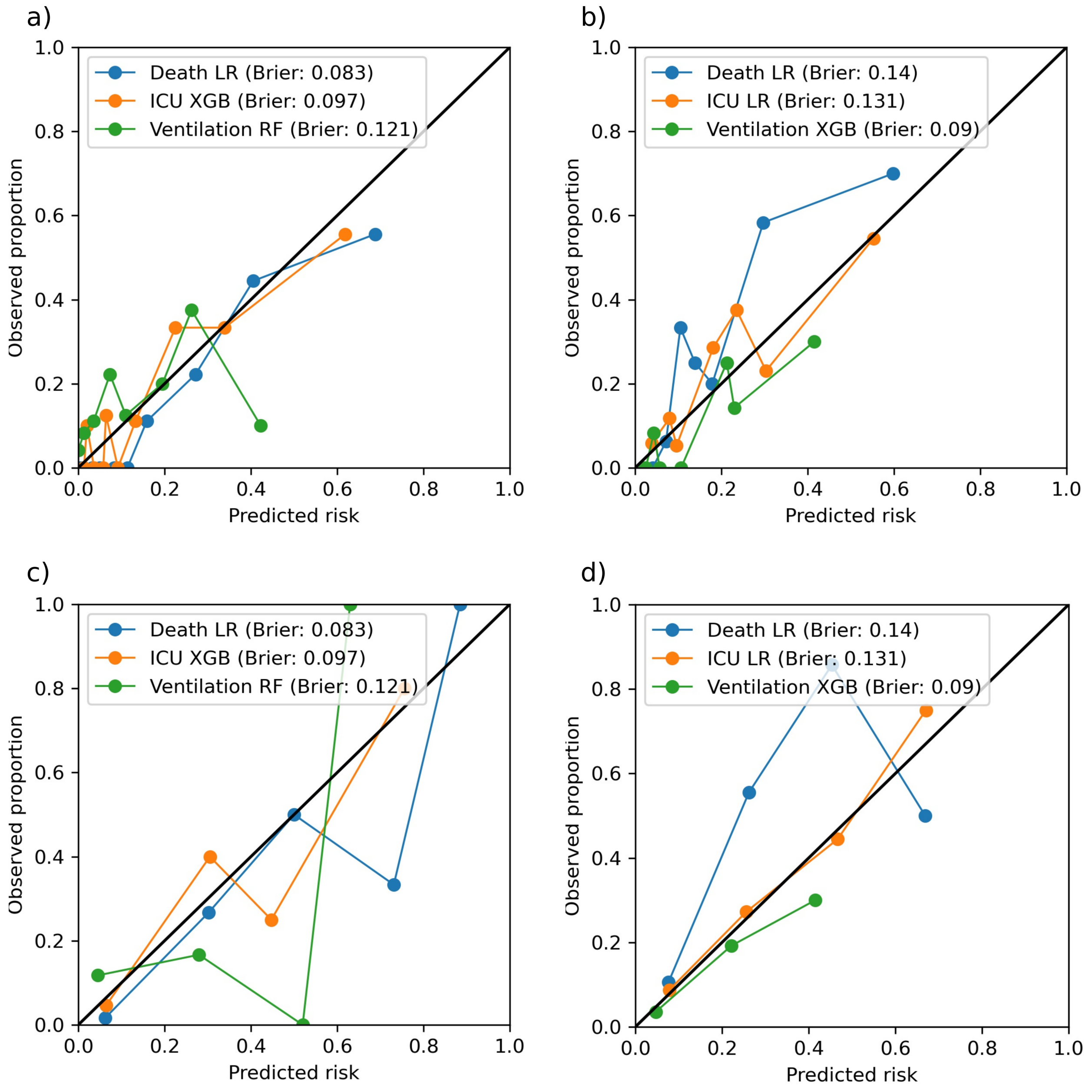

**Figure S4.** Calibration curves including brier scores of the best predictive machine-learning models **a)** based on numerical predictors using 10 bins, **b)** based on dichotomous predictors using 10 bins, **c)** based on numerical predictors using 5 equally spaced bins and **d)** based on dichotomous predictors using 5 equally spaced bins regarding the endpoints in-hospital mortality, transfer to ICU and mechanical ventilation.

#### *2.6 Additional ROC-curves using data of the first 48 hours*

We present ROC-Curves of our predictive machine-learning models applied to test data based on the averaged laboratory values of the first 48 hours, age and biological sex.

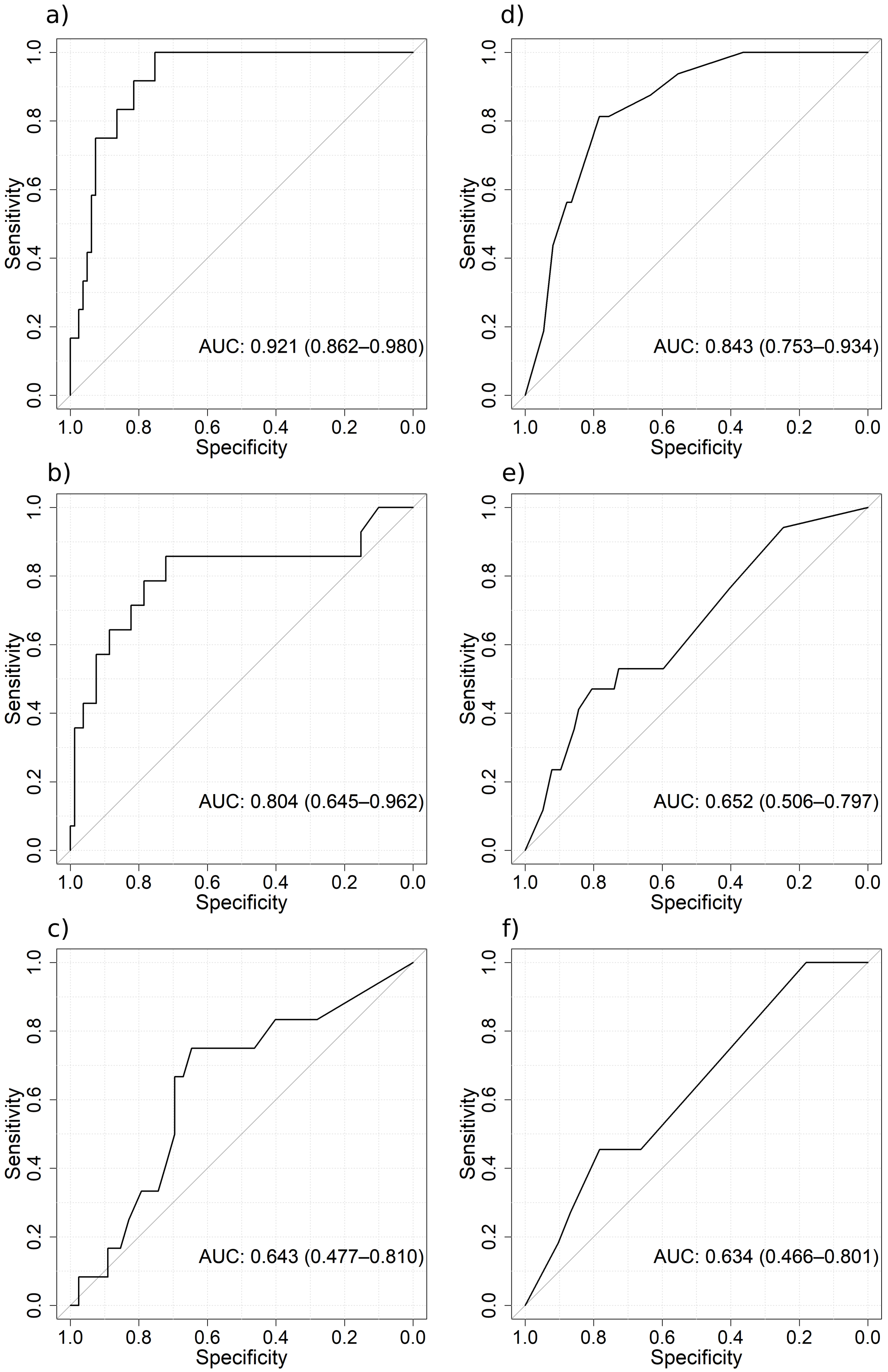
a)
AUC: 0.921 (0.862–0.980)
b)
AUC: 0.804 (0.645–0.962)
c)
AUC: 0.643 (0.477–0.810)
d)
AUC: 0.843 (0.753–0.934)
e)
AUC: 0.652 (0.506–0.797)
f)
AUC: 0.634 (0.466–0.801)
Sensitivity
Specificity
0.0
0.2
0.4
0.6
0.8
1.0

**Figure S5.** ROC-curves of the best predictive machine-learning models using test data based on the first 48 hours regarding the endpoints **a)** in-hospital mortality (Logistic Regression), **b)** transfer to ICU (XGBoost), and **c)** mechanical ventilation (Random Forest), **d)** in-hospital mortality (Logistic Regression with binarized covariates), **e)** transfer to ICU (Logistic Regression with binarized covariates), and **f)** mechanical ventilation (XGBoost with binarized covariates)

### *2.7 Hyperparameteroptimization*

To improve model performance we perform a hyperparameteroptimization for RandomForestClassifier and XGBoostClassifier. We use the Bayesian hyperparameter optimization function BayesSearchCV with a 5-fold cross validation[8]. We present the search space of the hyperparameters and the final hyperparameters in table S3 for RandomForestClassifier and in table S4 for XGBoostClassifier.

**Table S3**
List of choices for the hyperparameters of RandomForestClassifier used in the hyperparameteroptimization including the final configuration for the mechanical ventilation prediction model

| Hyperparameter | Seach space | Mechanical ventilation |
|---|---|---|
| Number of Trees | 50, 75, 100, 200, 300, 400, 500 | 100 |
| Max. depth of Trees | None, 2, 4, 6, 8, 10 | None |
| Min. number of samples required to be at a leaf node | 1, 2, 4 | 1 |
| Min. number of samples required to split an internal node | 2, 4, 6, 8, 10 | 2 |

**Table S4**
List of choices for the hyperparameters of XGBoostClassifier used in the hyperparameteroptimization including the final configuration for the ICU and dichotomized mechanical ventilation prediction model

| Hyperparameter | Search space | ICU | Mechanical ventilation |
|---|---|---|---|
| Number of Trees | 100, 200, 300, 400, 500 | 100 | 100 |
| Learning rate | 0.1 | 0.1 | 0.1 |
| Max. depth of Trees | 3, 4, 5, 6, 7, 8, 9, 10 | 4 | 6 |
| Min. sum of instance weight in a child | 1, 2, 3, 4, 5, 6 | 6 | 1 |
| Gamma | 0.0, 0.1, 0.2, 0.3 ,0.4 | 0.3 | 0 |
| Subsample ratio of training instances | 0.5, 0.6, 0.7, 0.8, 0.9, 1.0 | 0.9 | 1 |
| Colsample bytree | 0.5, 0.6, 0.7, 0.8, 0.9, 1.0 | 0.5 | 1 |

*2.8 Performance of all tested models on train data*

In this chapter we present the 5-fold cross-validated AUC of all mentioned predictive models on the training data with the chosen covariates. The best models are marked yellow.

**Numerical variables**

1) Logistic Regression

2) Logistic Regression with SMOTE

| Dead | Transfer to ICU | Mechanical Ventilation |
|---|---|---|
| 0.84<br>Urea, Age, CRP, Gluc | 0.79<br>Ca, CRP, Gluc, Age | 0.76<br>CRP, Ca |
| 0.806<br>'Age', 'CRP', 'Hst', 'Ca', 'BZ' | 0.84<br>'Ca', 'Gluc', 'PTT', 'Age', 'CRP' | 0.776<br>'Ca', 'Gluc', 'Sex', 'CRP' |

Random Forest

1) Variable Selection via Feature Importance

2) Using variables from logistic regression

3) Variable Selection via Feature Importance and SMOTE

4) Using variables from logistic regression and SMOTE

| Dead | Transfer to ICU | Mechanical Ventilation |
|---|---|---|
| 0.819<br>Age, CRP, Urea, GFR, Gluc, Ca | 0.782<br>Ca, CRP, Gluc, PTT, GOT | 0.778<br>Ca, CRP, Gluc |
| 0.815 | 0.764 | 0.777 |
| 0.83 Age, CRP, Hst, GFR, BZ, Ca | 0.815 Ca, CRP, BZ, PTT, GOT | 0.75 Ca, CRP, Gluc |
| 0.81 | 0.825 | 0.770 |

XGBoost

1) Variable Selection via Feature Importance

2) Using variables from logistic regression

3) Variable Selection via Feature Importance and SMOTE

4) Using variables from logistic regression and SMOTE

| Dead | Transfer to ICU | Mechanical Ventilation |
|---|---|---|
| 0.730<br>Age, Urea, PTT, Quick | 0.855<br>Age, Ca, CRP, Gluc, GOT | 0.741<br>Age, Ca, CRP, Gluc |
| 0.837 | 0.732 | 0.745 |
| 0.755 Age, Urea, PTT | 0.8<br>Age, Ca, CRP, Gluc, GOT | 0.69<br>Ca, CRP, Gluc |
| 0.8 | 0.76 | 0.773 |

**Categorical variables**

1) Logistic Regression

2) Logistic Regression and SMOTE

| Dead | Transfer to ICU | Mechanical Ventilation |
|---|---|---|
| 0.82<br>Urea, PTT, Age, GOT | 0.76<br>PTT, Ca, GOT, Sex | 0.68<br>Ca, PTT |
| 0.81<br>Hst, PTT, Age, RDW, GOT, GFR, Krea | 0.72<br>PTT,Ca,Sex | 0.76<br>GOT, Ca, Age, PTT, Sex, CRP |

Random Forest

1) Variable Selection via Feature Importance

2) Using variables from logistic regression

3) Variable Selection via Feature Importance and SMOTE

4) Using variables from logistic regression and SMOTE

| Dead | Transfer to ICU | Mechanical Ventilation |
|---|---|---|
| 0.754<br>Urea, GOT, PTT, Sex, RDW | 0.752<br>Ca, PTT, GOT | 0.708<br>Ca, PTT, CRP |
| 0.809 | 0.693 | 0.716 |
| 0.78<br>Urea, PTT, RDW | 0.74<br>Ca, PTT, GOT | 0.78 Ca, Sex, CRP, GOT |
| 0.801 | 0.7 | 0.73 |

XGBoost

1) Variable Selection via Feature Importance

2) Using variables from logistic regression

3) Variable Selection via Feature Importance and SMOTE

4) Using variables from logistic regression and SMOTE

| Dead | Transfer to ICU | Mechanical Ventilation |
|---|---|---|
| 0.805<br>CRP, Urea, Age | 0.695<br>PTT, GOT, CRP | 0.801<br>Ca, PTT, CRP, GOT |
| 0.8 | 0.698 | 0.716 |
| 0.807<br>CRP, Urea, Age | 0.697<br>CRP, GOT, PTT | 0.71<br>Ca, PTT, GOT, CRP |
| 0.8 | 0.7 | 0.72 |


[1] McKinney W. Data Structures for Statistical Computing in Python, Austin, Texas: 2010, p. 56–61. https://doi.org/10.25080/Majora-92bf1922-00a.
[2] Harris CR, Millman KJ, van der Walt SJ, Gommers R, Virtanen P, Cournapeau D, et al. Array programming with NumPy. Nature 2020;585:357–62. https://doi.org/10.1038/s41586-020-2649-2.
[3] Hunter JD. Matplotlib: A 2D Graphics Environment. Comput Sci Eng 2007;9:90–5. https://doi.org/10.1109/MCSE.2007.55.
[4] Waskom M. seaborn: statistical data visualization. JOSS 2021;6:3021. https://doi.org/10.21105/joss.03021.
[5] Virtanen P, Gommers R, Oliphant TE, Haberland M, Reddy T, Cournapeau D, et al. SciPy 1.0: fundamental algorithms for scientific computing in Python. Nat Methods 2020;17:261–72. https://doi.org/10.1038/s41592-019-0686-2.
[6] Seabold S, Perktold J. Statsmodels: Econometric and Statistical Modeling with Python, Austin, Texas: 2010, p. 92–6. https://doi.org/10.25080/Majora-92bf1922-011.
[7] Pedregosa F, Varoquaux G, Gramfort A, Michel V, Thirion B, Grisel O, et al. Scikit-learn: Machine Learning in Python 2018.
[8] Head T, MechCoder, Louppe G, Iaroslav Shcherbatyi, Fcharras, Zé Vinícius, et al. Scikit-Optimize/Scikit-Optimize: V0.5.2. Zenodo; 2018. https://doi.org/10.5281/ZENODO.1207017.
[9] Chen T, Guestrin C. XGBoost: A Scalable Tree Boosting System. Proceedings of the 22nd ACM SIGKDD International Conference on Knowledge Discovery and Data Mining, San Francisco California USA: ACM; 2016, p. 785–94. https://doi.org/10.1145/2939672.2939785.
[10] Wickham H, Averick M, Bryan J, Chang W, McGowan L, François R, et al. Welcome to the Tidyverse. JOSS 2019;4:1686. https://doi.org/10.21105/joss.01686.
[11] Robin X, Turck N, Hainard A, Tiberti N, Lisacek F, Sanchez J-C, et al. pROC: an open-source package for R and S+ to analyze and compare ROC curves. BMC Bioinformatics 2011;12:77. https://doi.org/10.1186/1471-2105-12-77.
[12] Flier FJ, Hirs WM. The challenge of an international classification of procedures in medicine. Medinfo 1995;8 Pt 1:121–5.
[13] International classification of procedures in medicine. 1: Procedures for medical diagnosis, laboratory procedures, preventive procedures, surgical procedures, other therapeutic procedures, ancillary procedures. Geneva: World Health Organization; 1978.
[14] Youden WJ. Index for rating diagnostic tests. Cancer 1950;3:32–5. https://doi.org/10.1002/1097-0142.
